# Optimized DBS in pediatric dystonia restores balance in transmission of signals within pallidothalamic network by modulating neural oscillations in deep brain regions

**DOI:** 10.1101/2025.10.07.25337522

**Authors:** Maral Kasiri, Mehrnaz Asadi, Jaya Nataraj, Sumiko Abe, Terence D. Sanger

## Abstract

Deep brain stimulation (DBS) is a neuromodulation technique commonly used for treatment of movement disorders, including dystonia. Stimulation of the globus pallidus internus (GPi) of basal ganglia or the subthalamic nucleus (STN) typically confers clinical benefit, although the specific mechanism of action remains unclear. Previous studies in dystonic patients show abnormalities in low-frequency activity in GPi and other motor sensory regions such as STN, ventralis oralis anterior/posterior (VoaVop), and ventral anterior (VA) nuclei of thalamus. We hypothesize that DBS works in part by modulating transmission of abnormal signals in low frequency bands between different brain regions, both at the stimulation site (e.g. GPi) and distant deep brain regions.

To test this hypothesis, we used a novel transfer function analysis that has not previously been utilized to study neural signal transmission. We recorded intracranial signals from 13 pediatric and young adult patients with dystonia, with and without stimulation. We performed transfer function analysis to compare the mean transfer function gain—representing signal amplification from input to output—across deep brain pathways in low-frequency bands, under both DBS-on and DBS-off conditions. Our results show that DBS modulates signal transmission between different brain regions. In particular, GPi stimulation increased transfer function gains from pallidum to thalamic motor subnuclei, especially in the beta and gamma frequency bands. These findings support the hypothesis that DBS decreases inhibitory output from GPi to thalamus through enhanced high-frequency transmission, offering insight into its mechanism of action. This, in turn, may provide fundamental knowledge for the development of closed-loop DBS, particularly in controlling the intensity and pattern of stimulation. A better understanding of neuromodulation could also help to further the design of brain-computer interfaces and neurorehabilitation systems.

## 1 Introduction

Deep brain stimulation (DBS) is a neuromodulatory intervention that delivers electrical impulses to specific brain regions to treat a range of movement and neurological disorders [1], including Parkinson’s disease, dystonia [2], essential tremor [3], epilepsy [4], and Alzheimer’s disease [5], as well as psychiatric disorders such as obsessive compulsive disorder [6] and major depression disorder [7]. DBS is commonly thought to act by modulating or blocking activity within specific brain regions. Here, we explore the possibility that DBS may also act by modulating the transmission of signals between those regions. In particular, we examine whether DBS alters the transmission of low-frequency oscillations between basal ganglia and thalamic regions, in children with primary and secondary dystonia [2, 8–10]. “Dystonia is defined as a movement disorder in which involuntary sustained or intermittent muscle contractions cause twisting and repetitive movements, abnormal postures, or both” [11, 12]. The underlying mechanism of dystonia is not known [2, 11]. However, previous research suggests several possible origins of dystonic motor symptoms, including decreased focus on intended patterns of muscle activity [13], imbalances between midbrain and striatal excitatory/inhibitory signaling [14], abnormal patterns of subcortical activity [15, 16], and excessive gain in basal ganglia or peripheral loops [15, 17, 18].

Mathematical models of dystonia propose that the associated motor abnormalities may result from an imbalance between the direct and indirect pathways of basal ganglia circuitry. Specifically, an increased gain in the indirect pathway relative to the direct pathway could lead to unstable and uncontrolled synchronous oscillations within the cortex and basal ganglia, potentially impairing motor control [18]. Such oscillatory behavior is assumed to contribute to dystonia, as effective motor control necessitates a controlled balance between these two pathways, as depicted in Fig. 1 (a). Moreover, recordings from our previous works suggest that these abnormal patterns are associated with increased activity within globus pallidus internus (GPi)—a primary inhibitory nucleus of the basal ganglia—during voluntary movement [18], particularly at low frequencies in the theta (4-8 Hz) and alpha (8-13 Hz) frequency bands [15, 16], providing support for these mathematical models. Therefore, excessive inhibitory output from the GPi to the thalamus could disrupt the normal excitation-inhibition balance in the thalamo-cortical networks. This results in both insufficient inhibition of undesired movement and insufficient disinhibition of intended movement [15].

**Fig. 1.**
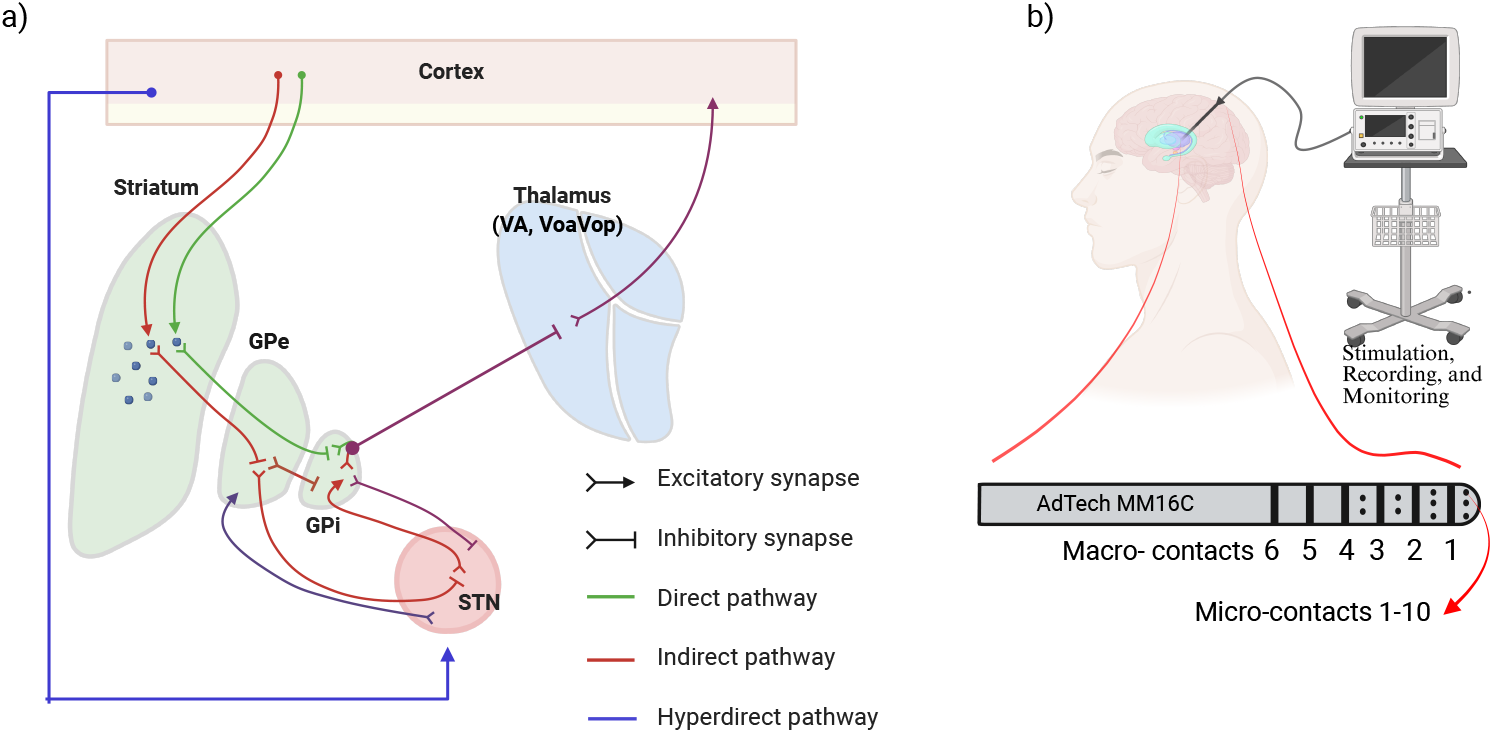
a) Schematic representation of the direct and indirect pathways in the cortico-basal ganglia-thalamo-cortical loop. This diagram illustrates the neural pathways within the basal ganglia, thalamus, and cortex, emphasizing their roles in the facilitation and inhibition of movement. The direct pathway (shown in green) promotes the activation of motor cortex through a series of disinhibitory signals. Conversely, the indirect pathway (depicted in red) inhibits movement by suppressing motor cortex activity via inhibitory signalling through thalamic nuclei. The co-activation of these two pathways could be essential for movement initiation and accurate motor control. b) Schematic of an externalized sEEG lead in a deep brain region. This figure illustrates the positioning of the lead within a specific deep brain area, connected to a monitoring unit capable of stimulating and recording simultaneously [31].

GPi or subthalamic nucleus (STN) DBS at clinically effective frequencies, 120–190 Hz for GPi [19] and 130–185 Hz for STN [20], often provides significant therapeutic benefit [2, 9, 10]. Moreover, in our previous study, we used transfer function modeling to predict high-frequency evoked responses to DBS and showed that stimulation pulses are more likely to propagate through the same pathways as intrinsic neural signals [17]. However, the low-frequency components of these transfer functions, which represent the underlying neural transmission, were not studied. Therefore, how DBS pulses influence the intrinsic signals is still unclear. Possible explanations are facilitating, blocking, or changing the power and transmission delays in several frequency bands, or combinations of these effects [17]. Understanding how DBS signals flow through deep brain networks and influence motor circuitry, especially pathways from pallidum to thalamic motor nuclei, would enhance our understanding of both the pathophysiology of dystonia and the mechanisms of DBS in the treatment of dystonia.

Transfer functions are commonly used in engineering to analyze transmission of band-limited information between a source and destination. Here, we apply this method of analysis to examine the amplification of neural signal transmission in multiple frequency bands between different brain regions and to investigate how this transmission changes in the presence of DBS with various stimulation frequencies. We hypothesize that stimulation of GPi and STN modulates the transmission of pathological low-frequency neural activity within the indirect pathway, particularly from STN to GPi. Additionally, we investigated whether GPi and STN stimulation influences low-frequency activity propagation through the direct and indirect pathway outputs, specifically from the GPi to thalamic nuclei, including the ventral oralis anterior/posterior (VoaVop) and ventral anterior (VA) nuclei. Our analysis focused on standard low-frequency bands (theta: 4–8 Hz, alpha: 8–12 Hz, beta: 13–30 Hz, and low gamma: 30–50 Hz) to characterize these effects. Furthermore, we examined how varying stimulation frequencies impact signal transmission patterns across three key anatomical pathways in the pallidothalamic network: STN to GPi, GPi to VoaVop, and GPi to VA.

To study the effects of DBS, neural signals were acquired using externalized stereoelectroencephalography (sEEG) leads implanted in basal ganglia and thalamic nuclei of 13 children and young adults with childhood-onset dystonia of various underlying etiologies (genetic, acquired, or metabolic). Recordings were obtained in both DBS-on and DBS-off conditions. Empirical transfer functions and their magnitude responses, up to 50 Hz, were then estimated for pathways connecting each pair of these motor regions, in both conditions. Together, multi-electrode recordings and transfer function analysis offer valuable insight to understand the transmission of neural information between regions. This method enables estimation of information flow at low frequencies—reflecting intrinsic neural activity—even during active DBS, as clinical stimulation frequencies are typically above 50 Hz. To our knowledge, this analytical approach has not previously been used in studies of DBS effects on signal transmission. Thus, it offers a novel framework to investigate how DBS modulates neural communication within deep brain circuits, potentially providing critical insights into the mechanisms underlying DBS efficacy. Such insights could inform future improvements in DBS programming protocols and the development of closed-loop DBS systems.

## 2 Materials and Methods

### 2.1 Subjects

Thirteen pediatric and young adult patients undergoing deep brain stimulation (DBS) surgery for the treatment of dystonia participated in this study (1). Patients were previously diagnosed with dystonia by a pediatric movement disorder physician (T.D.S.) using standard criteria [9, 11]. All patients had a prior history of insufficient response to both botulinum toxin injections and standard pharmacotherapy. All patients or parents of minors provided signed informed consent for surgical procedures in accordance with standard hospital practice at Children’s Health Orange County (CHOC) or Children’s Hospital Los Angeles (CHLA). The patients or parents of minor patients also consented or assented to Health Insurance Portability and Accountability Act (HIPAA) authorization for the research use of protected health information and the recorded electrophysiological data.

**Table 1.**
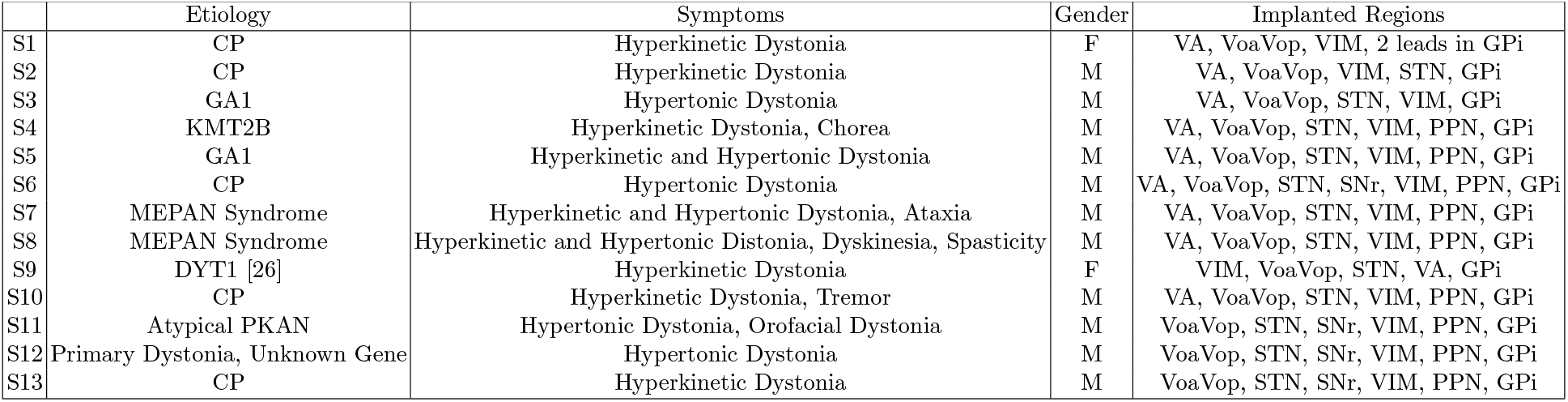
Patient demographics. CP: Cerebral Palsy; GA1 [21]: Glutaric aciduria type 1; KMT2B: Lysine Methyltransferase-2B [22]; MEPAN [23, 24]: Mitochondrial Enoyl CoA Reductase Protein-Associated Neurodegeneration; PKAN [25]: Pantothenate Kinase-Associated Neurodegeneration; F: female; M: male.

### 2.2 Surgical Procedure

As part of a previously described clinical procedure for determining DBS targets [27, 28], up to 12 temporary AdTech MM16C (Adtech Medical Instrment Corp., Oak Creek, WI, USA) externalized sEEG depth electrodes were implanted into potential DBS targets using standard stereotactic procedure [2, 27]. Based on prior studies of DBS in children with dystonia, typical targets include subthalamic nucleus (STN) and globus pallidus internus (GPi) in basal ganglia; ventral intermediate (VIM), venral oralis anterior and posterior (VoaVop), and ventral anterior (VA) nuclei of thalamus [2, 8]; and substantia nigra reticulata (SNr) [29]. A schematic of an externalized lead and the targeted deep brain regions are shown in Fig. 1 (b). Targeting of thalamic subnuclei was confirmed using median nerve stimulation, to which various thalamic nuclei are known to have greater or lesser evoked response [30]. Electrode locations were later confirmed by coregistration of the preoperative magnetic resonance imaging (MRI) with postoperative computed tomography (CT) scan [16].

The patients were then transferred to the inpatient neuromodulation monitoring unit (NMU) for stimulation pattern testing in order to identify the optimal targets for permanent DBS placement [27]. During this phase, lasting up to 7 days, local field potential (LFP) signals were captured via depth electrodes with and without stimulation. After these 7 days, the optimal stimulation target(s) were identified, and the temporary depth leads were then removed. At a minimum of two weeks after the removal of the temporary leads, permanent leads were implanted at the chosen DBS targets. After two weeks following placement of permanent electrodes, these electrodes were connected to pulse generators placed in the chest [27].

### 2.3 Instrumentation and Setup

Elecrophysiological recordings were acquired starting at 24 hours after clinical implantation of the temporary depth leads and after localization of the macro-electrode contacts on each lead using the co-registered imaging studies. Each implanted lead contains 6 low-impedance (1–2 *kΩ*) ring macro-electrode contacts with 2 mm height and 5 mm on-center spacing, as well as 10 high-impedance (70–90 *kΩ*) microwire electrodes (50- *µm* diameter; micro-contacts). The micro-contacts are arranged in groups of 2 or 3, evenly spaced around the circumference of the lead shaft, halfway between neighboring pairs of the first four macro-contacts. A schematic of the electrode is shown in Figure. 1 (b). The leads were connected to Adtech Cabrio− connectors which include a custom unity-gain preamplifier for each micro-contact for reduction of noise and motion artifacts. Macro-contacts bypassed the preamplifiers enabling external electrical stimulation. This setup allows clinicians to stimulate through macro-contacts while recording from all the micro- and macro-contacts simultaneously. In this study, we used the data recorded from the high-impedance electrodes (micro-contacts). These signals were amplified and sampled at 24 kHz by a custom recording setup that includes a PZ5M 256-channel digitizer and RZ2 processor and stored in a RS4 high speed data storage device (Tucker-Davis Technologies Inc., Alachua, FL, USA) [16].

### 2.4 Electrophysiological Recordings

We recorded intracranial LFPs from all patients, with and without stimulation, following our standard clinical testing protocol [27]. For this study, we analyzed data from electrodes in GPi, VoaVop, and STN, and VA. The decision to limit our analysis to this subset of micro-electrode recordings was made because of the key involvement of these structures in the direct and indirect pathway of the basal-ganglia-thalamo-cortical motor circuitry, affecting motor symptoms. The recording and stimulation regions were confirmed by the post-operative analysis of MRI and CT scans, as explained in the previous section.

#### Baseline recording

Baseline data were recorded from all micro-contacts and all leads while patients were awake and sitting comfortably in bed. In all cases, dystonia was present during movement as evident from the presence of dystonic posturing.

#### Recording during DBS

Stimulation pulses of 90 *µs* pulse width and 3 *V* amplitude were administered at four different frequencies (55, 85, 185, and 250 Hz), through two adjacent macro-contacts (anode and cathode) at a time, ensuring that the cathode (negatively charged electrode) is in the desired DBS target. For stimulation frequencies above 60 Hz, the stimulation protocol was altered to include voltage ramping from 0 to 3 V at a rate of 4 seconds per volt, for the comfort of the patient [2]. We administered approximately 1200 pulses per stimulation frequency after voltage ramping was complete. The LFP signals were recorded through the micro-contacts simultaneously during the stimulation.

### 2.5 Data treatment and filtering

All data was notch filtered at 60 Hz and its first two harmonics. A 4^*th*^ order butterworth high-pass filter was applied with a cutoff frequency of 1 Hz to remove drift. The monopolar recordings from the micro-contacts on each row of the lead (total of 10 per lead) were re-referenced to a bipolar montage by computing the voltage difference for each pair of adjacent micro-contacts. Re-referencing produced 8 channel recordings per lead, as shown in Figure. 2 (b). This process removes the common mode noise and uncovers the underlying neural activity [15]. It is important to note that a post-processing step to remove stimulation artifacts is not necessary in this case, because all stimulation artifacts will occur at multiples of the stimulation frequency and therefore are effectively removed by low-pass filtering below 50 Hz. For consistency throughout our analyses, we used a maximum of 10 seconds of signals per stimulation setting and/or baseline recording.

**Fig. 2.**
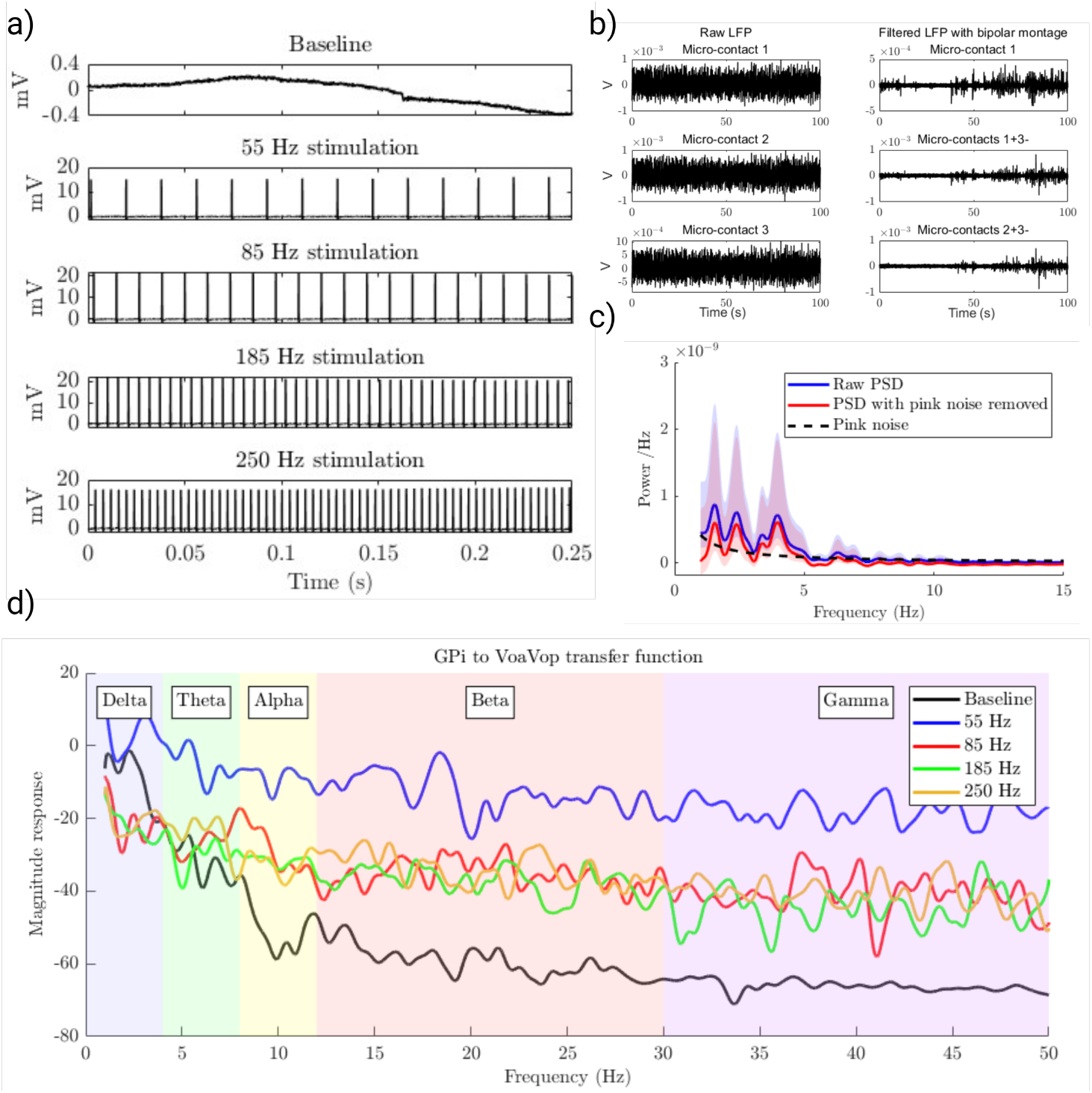
a) Sample of raw data recorded over 0.25 seconds during baseline, and during DBS at 55, 85, 185, and 250 Hz, demonstrating contamination from stimulation artifacts; b) Baseline LFP recordings from three micro contacts on the first row of an STN lead (left). The same baseline LFP recordings after re-referencing to a bipolar montage and high-pass filtering (right); c) Raw Power Spectral Density (PSD; blue), the fitted pink noise (black dashed line), and the PSD after removal of the pink noise component (red); d) This figure depicts samples of GPi to VoaVop transfer function magnitudes during baseline and during GPi-DBS at 55, 85, 185, 250 Hz stimulation frequencies, highlighting a significant increase in magnitude response across theta, alpha, beta, and low gamma bands, while stimulation is on.

### 2.6 Computation of transfer function

An empirical transfer function can be computed using several different methods. The magnitude of the transfer function is a linear measure of the amount of information transferred between two points at each individual frequency. In this study, in particular, we wanted to evaluate the changes in the magnitude response of the transfer function representation of each pathway connecting two points between two electrodes. The transfer function between each two points is computed as the ratio of the output Fourier transform to the input Fourier transform [17, 32, 33]. The power spectral density (PSD) can be estimated as the value of the Fourier transform to the power of two, divided by two times the frequency spacing. Therefore, we can estimate the transfer function magnitude by:

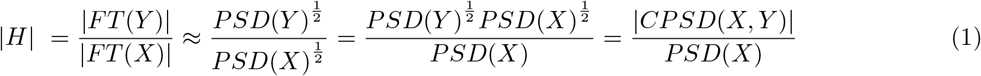

Where *PSD*(*X*) is the power spectral density of the input and *CPSD*(*X, Y* ) is the cross power spectral density between input and output. This single-input-single-output (SISO) transfer function model is a linear representation of each pathway, all of which can be combined to comprise a multiple-input-multiple-output (MIMO) system. However, here, we analyze each pathway individually, therefore *H* represents the pathway transfer function (*H*(*ω*)) itself [34]. This model can characterize how each region affects the signal transmission within the whole network and how that network responds to exogenous stimulation (DBS) [34]. It is worthy to note that transfer functions and coherence measure different aspects of signal relationships. Coherence quantifies the linear correlation between two signals at each frequency, reflecting how consistently they co-vary in both amplitude and phase. In contrast, transfer functions describe how input signals are transformed into output signals, capturing both magnitude and phase information. Our analysis specifically focuses on the magnitude component |*H*(*f* )|, which reflects the strength of signal transmission at each frequency. Under the assumption of a linear time-invariant (LTI) system with unit coherence, the transfer function magnitude serves as a reliable estimator of the relationships between two signals. We assume unit coherence between the signals, and to compensate for this assumption, we add a regularization parameter to account for inputs beyond the primary input node. In addition, in the process of deriving transfer functions from PSDs, the incorporation of a regularization constant, *ϵ*, in the denominator is a critical step to ensure numerical stability and accuracy. This need arises particularly at frequencies where the PSD of the input approaches zero, leading to potential numerical instability due to division by values close to zero. To mitigate this, a small value (*ϵ*) is added to the denominator, effectively ensuring that the transfer function remains well-defined across all frequencies [35]. The value of *ϵ* was determined to be 5*e−* 11 through a trial-and-error approach, aiming to find the smallest constant that prevents numerical issues while minimally impacting the fidelity of the transfer function. The rationale behind adding a constant to the denominator is due to the assumption that any other inputs contributing to the output of the system follow a constant distribution, implying that the system’s response at frequencies where the input PSD is very low is dominated by these other inputs. Consequently, the introduction of *ϵ* can be viewed as a representation of the baseline level of these other contributions, providing a more realistic and stable characterization of the system’s behavior. This approach, while empirical, aligns with common practices in system identification and signal processing, where balancing the fidelity of the model with the practical limitations of real-world data is often necessary [35, 36]. Thus, the new transfer function equation is:

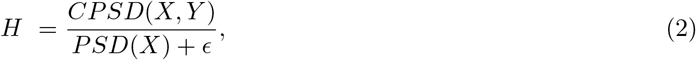

where *ϵ* is the regularization constant.

For each subject, the PSD for each channel and the pairwise cross spectral density (CPSD) between the channels were computed in the DBS-on (separately for different frequencies) and DBS-off conditions using Welch’s method with a *∼* 3 *s* hamming window, 30% overlap, and over a frequency range of 1 to 50 Hz. The computed PSDs and CPSDs are contaminated with pink noise which is characterized by a fractional 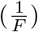 decay. Therefore, we fitted a function, 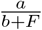, to each computed spectra and then subtracted that fractional trend from the data, as illustrated in Figure. 2 (c). Once pink noise was removed, the transfer function model of each pathway was constructed using Equation 2, during baseline and DBS-on, for all recording pairs.

The aim of this study is to assess the effect of different frequencies of stimulation on the transmission of low frequency oscillations (LFO) within deep brain networks. Due to the wide usage of low frequency bands in neurology, we analyze these effects in EEG standard frequency bands (theta [4-8 Hz], alpha [8-13 Hz], beta [13-30 Hz], and low gamma [30-50 Hz]), as depicted in Figure. 2 (d). Therefore, the mean transfer function gains were computed at each of these frequency bands and the effect of stimulation at different frequencies was compared within each frequency band. Note that the frequencies analyzed for the transfer function analysis (0.5-50Hz) are considerably lower than the stimulation frequencies (55-250 Hz), so the changes we analyze are likely to represent neural effects of stimulation rather than a direct electrical stimulation or blocking effect.

### 2.7 DBS electrode localization

Preoperative anatomical brain scans were acquired using a MAGNETOM 3T MRI scanner (SIEMENS Medical System, Erlangen, Germany). A magnetization-prepared rapid gradient-echo (MP RAGE) sequence was used to achieve precise anatomical mapping, with repetition time (TR) of 1800 ms, an echo time (TE) of 2.25 ms, a flip angle of 8^°^, a voxel size of 1 *mm*^3^, and a field of view (FOV) of 240 *mm*^2^. After the operation, computed tomography (CT) scans were acquired with GE (GENERAL ELECTRIC Healthcare, Milwaukee, WI, USA) CT scanner.

Brain regions were identified and segmented using the Freesurfer software [37] and localized deep brain regions. Segmentation was then manually adjusted by FreeView (a view tool of FreeSurfer), a transformation matrix of deep brain regions (including thalamus, pallidum, and brainstem) was computed to convert from the spatial coordinates of each individual’s regions to Montreal Neurological Institute (MNI) space using the FSL FLIRT tool [38, 39]. Thereafter, the transformation matrices were used to transform the sub-regions from the DISTAL atlas (built by LEAD-DBS [40]) to the “actual” individual space.

For visualization of lead trajectories, standard software cannot be used [41] because they do not support the specifications of our sEEG leads and are unable to visualize 10 leads simultaneously. Therefore, for localization of sEEG leads, we used the metal artifacts generated by macro-contacts in the CT-scans to create a binary mask of spatial coordinates of the macro-contacts for all Adtech sEEG leads [42]. The CT and T1-volumes were aligned using the FLIRT tool in FSL (Fig.3 (a) and (b)). 3D voxel values were then projected onto 2D images [43] to precisely localize the lead trajectories.

After the lead trajectories were estimated based on the projection coordination (Figure 3), a linear calculation was applied to find the location of micro-contacts, which are located between macro contacts based on the device specification. Finally, the DSI-studio package [44] was used to integrate the sEEG leads coordinates with the atlas images as depicted in 3 (c). This method enabled us to localize each row of microcontacts on the sEEG leads, which is important for target discrimination during stimulation testing as some leads target multiple deep brain regions.

**Fig. 3.**
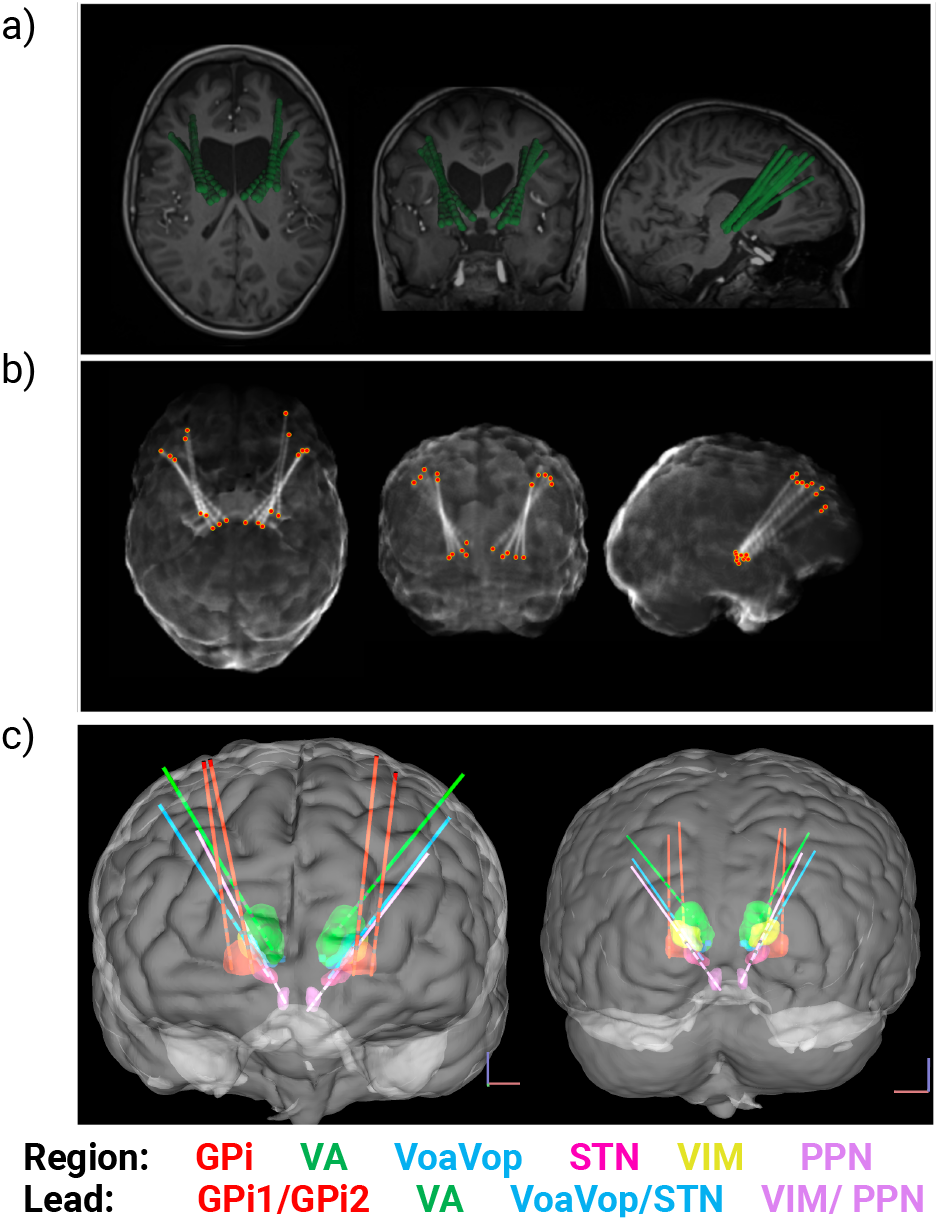
(a,b) sEEG lead trajectory reconstruction in axial (left), coronal (middle), and sagittal (right) views in an individual space. (a) The CT scan is marked as a dense structure and fitted onto the structural images. (b) Projection images in axial, coronal, and sagittal views are used to reconstruct the positions of the sEEG lead electrodes. (c) Anterior (left) and posterior (right) views of reconstructed lead trajectories and nuclei segments in one patient; DBS targets were segmented and the spatial coordinates of the leads were calculated. The figure illustrates GPi (red), VA (green), VoaVop (blue), VIM (yellow), STN (magenta), and PPN (violet) along with the reconstructed sEEG lead trajectories using DSI studio package.

### 2.8 Statistical analysis

Initial visual inspection was done on every patient’s data, exploring if there is a consistent pattern within the transfer function gains with the stimulation frequencies and the stimulation location. After visual inspection, we identified an overall increase in the transfer function gains. Linear mixed-effects model analysis was conducted on the average gains at each frequency band to estimate their relationship with the stimulation frequencies, using the lme4 and emmeans packages in R studio [45, 46].

For additional analysis, we separated the stimulation data into two groups: 1. The stimulation setting (frequency and stimulation macro-contact pairs) was not clinically effective. 2. The stimulation setting (frequency and stimulation macro-contact pairs) was clinically effective. The same analysis was done to compare if the transmission of signals were affected differently during the clinically effective stimulation setting, compared to the ineffective setting.

## 3 Results

We fitted linear mixed-effects models to the mean transfer function gains at each frequency band to estimate the effects of stimulation, including the impact of various stimulation frequencies, on these gains. After careful analysis and visualization of the data, we observed that the relationship between stimulation frequency and transfer function gain was not monotonic or linear. Therefore, we treated the stimulation frequency as a discrete variable and we performed pairwise comparisons to assess the significance of the observed changes relative to the baseline transfer function gains, across the standard EEG frequency bands, and compared how they differ from each other. All reported p-values of multiple comparisons are Bonferroni-corrected for increased reliability of the results. The fitted nested linear mixed effect model in R to assess the effect of DBS frequency on different pathways’ signal transmission was defined as:

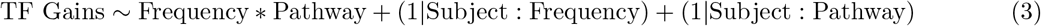

where subjects are the random effects, to account for individual variations. The stimulation frequency and the pathways served as both fixed and random effects nested within each subject in the models, contributing to the overall variance in the data and the within-subject intercept variability. We then performed a pairwise comparison between the changes of transfer function gains for all standard frequency bands during STN stimulation 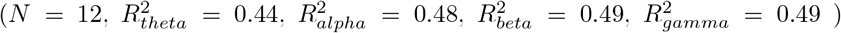 and during GPi Stimulation 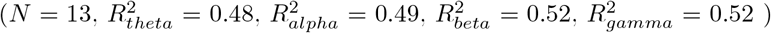 . We excluded the delta band due to contamination at the low frequency filter cutoff. One subject did not have leads implanted in STN, and was excluded from STN stimulation analysis. The significance of the estimated coefficients was tested by type III Analysis of Variance Table with Satterthwaite’s method and the p-values were computed from the test statistic based on chi-squared distribution with threshold of 0.05. The results from the model show that stimulation in both STN and GPi increased transfer function gains significantly in all routes for all patients, regardless of the frequency bands as depicted in Figure 4 and Figure 7.

**Fig. 4.**
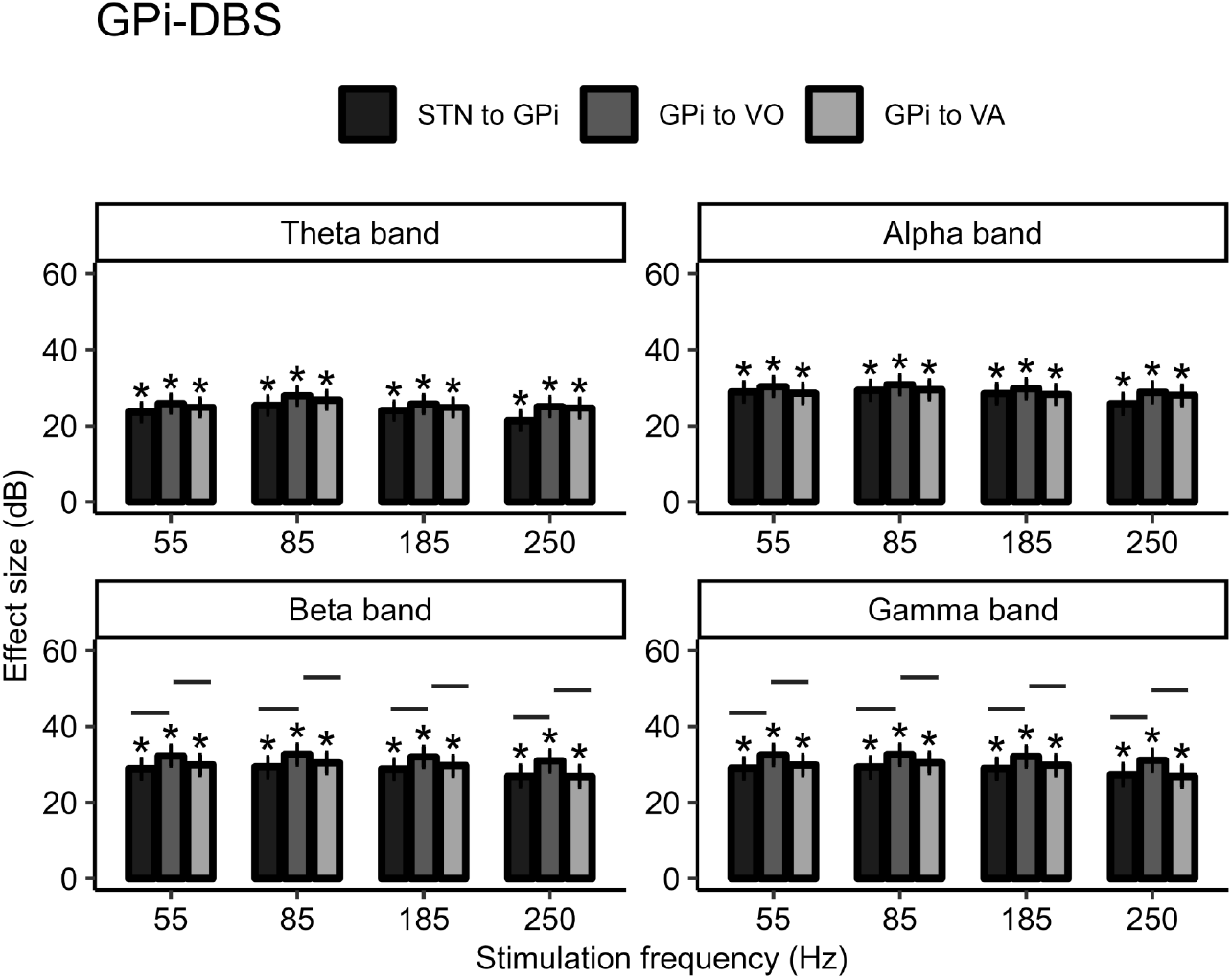
Statistical results: Effect of GPi-DBS on the transfer function gains within the pathways, with respect to the baseline transfer function gains. The lines indicate the significance within pathways and the *s indicate the significance of each bar with respect to baseline gains (DBS-off). Transfer function gains of theta, alpha, beta, and gamma bands at the baseline activity and during 55, 85, 185, 250 Hz GPi-DBS and STN-DBS. All the transfer function gains during DBS are significantly larger than the gains during the baseline (DBS-off), illustraded with *.

We then performed a pairwise multiple comparison using Kenward-Roger’s F-test with the emmeans [46] package to find the differences between each pathway (GPi-VoaVop, GPi-VA, and STN-GPi) and their interaction (conditional effect) with stimulation frequencies (0 [DBS off], 55, 85, 185, and 250 Hz). This method compares the estimated marginal means to discover the effect of stimulation frequency on the patterns of transmission.

### 3.1 GPi stimulation

#### Theta and Alpha bands

The results of pairwise comparison showed no significant changes within pathways at frequencies of stimulation.

#### Beta band

The results of pairwise comparison with respect to baseline showed that during all stimulation frequencies 55, 85, 185, and 250 Hz GPi-DBS, GPi-VoaVop beta band gains are significantly higher than that of STN-GPi (Estimates _Beta_ = 2.59, 2.42, 2.38, 3.14 respectively: *p − value <* 0.01) ^5^ and GPi-VA (Estimates _Beta_ = 1.99, 1.82, 1.92, 3.70: *p − value <* 0.01), while these gains are not significantly different at baseline activity.

#### Gamma band

Similar to the results at Beta band, the results of pairwise comparison with respect to baseline showed that during all stimulation frequencies 55, 85, 185, and 250 Hz GPi-DBS, GPi-VoaVop gamma band gains are significantly higher than that of STN-GPi (Estimates _Gamma_ = 2.58, = 2.37, 2.25, 2.88 respectively: *p − value <* 0.01) and GPi-VA (Estimates _Gamma_ = 2.11, 1.59, 1.72, 3.70: *p − value <* 0.01), while these gains are not significantly different at baseline activity.

In summary, as shown in Figure 4, the transfer function gains did not differ within the stimulation frequencies, especially the clinically effective stimulation frequency, 185 Hz. However, the results show that during all stimulation frequencies, GPi-VoaVop gains increased significantly compared to the GPi-STN and GPi-VA gains at beta and gamma bands, but not the theta and alpha bands. Moreover, the statistical analysis on the effect of GPi-DBS on the transfer function gains at each frequency band is shown in Figure 5. These results illustrate that the GPi-DBS has the lowest effect size (least increase) on the theta band transmission and the highest at beta and gamma band transmission. This indicates that the DBS facilitates transmission of beta and gamma activity more than the transmission of theta activity in the pallido-thalamic network.

**Fig. 5.**
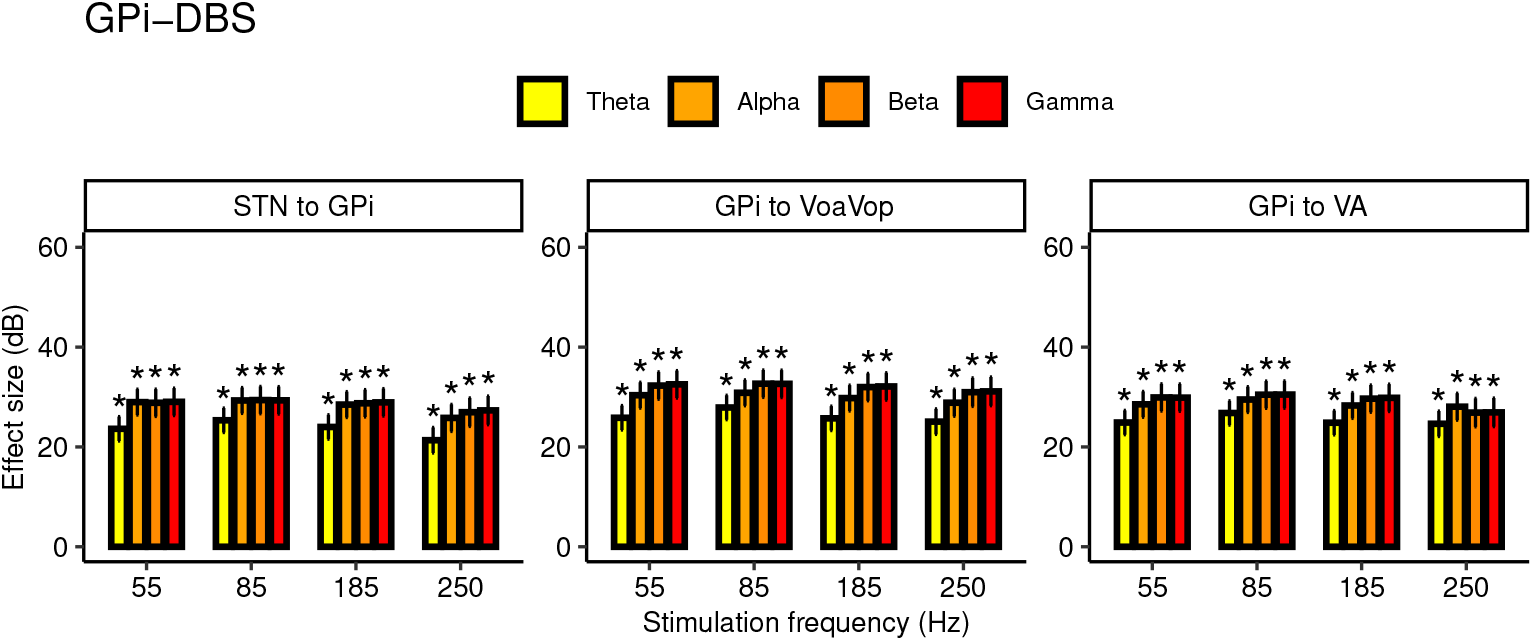
Statistical results: Effect of GPi-DBS on the transfer function gains within the frequency bands for each pathway.

#### Clinically optimal versus non-optimal GPi-DBS setting

We marked the optimal stimulation settings (stimulation macro-contacts, leads, and stimulation frequencies) for those patients that responded to GPi-DBS and performed the same statistical analysis. All 13 patients responded to GPi DBS either at 185 or 55 Hz. We fitted a linear mixed effect model as:

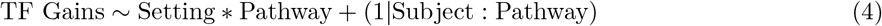

where “setting” stands for the DBS setting regardless of the stimulation frequency and has three levels: 1. DBS-off, 2. non-optimal setting, 3. optimal setting. After we fitted the models for each frequency band gain 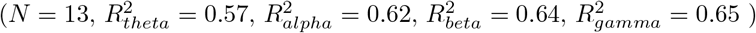, we performed a pairwise comparison to determine whether the “optimal” and “non-optimal” settings had different effects on the transmission of signals within deep brain regions. Figure 6 shows the transfer function gains across all frequency bands (theta, alpha, beta, gamma) and pathways. Each color represents one patient with their mean gain and standard deviation.

**Fig. 6.**
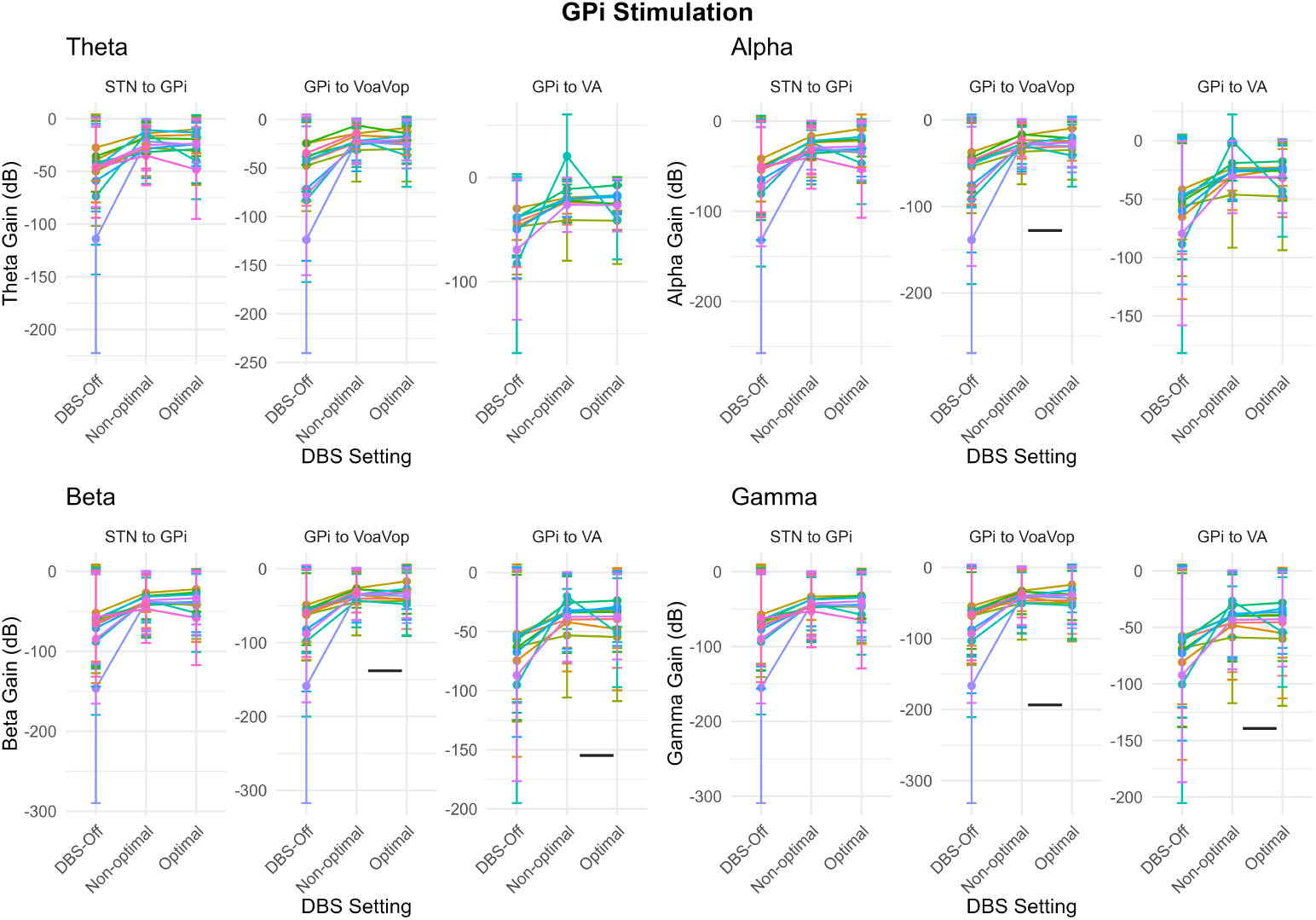
Effect of optimal versus non-optimal GPi-DBS setting of transfer function gains, shown for each pathway and frequency band, for all patients. Here, GPi DBS was effective clinically for all patients.

The results showed that there is no difference between the effect of optimal versus non-optimal DBS on the STN to GPi transfer function gains, as shown in Figure 6. However, ANOVA testing revealed that the GPi-VO gain during the optimal DBS is significantly larger than that of non-optimal DBS, in alpha (*Estimate* = 2.37; *p − value <* 0.01), beta (*Estimate* = 2.69; *p − value <* 0.01), and gamma (*Estimate* = 2.95; *p−value <* 0.01) bands. A similar effect was seen in the GPi to VA transfer function gains in only beta (*Estimate* = 1.28; *p − value <* 0.05) and gamma (*Estimate* = 1.32; *p − value <* 0.05) bands. For simplicity of the figure we are not showing the significance between the DBS-off versus optimal and non-optimal stimulation setting.

### 3.2 STN stimulation

#### Theta band

The results of pairwise comparison showed no significant changes within pathways at frequencies of stimulation (Figure 7).

**Fig. 7.**
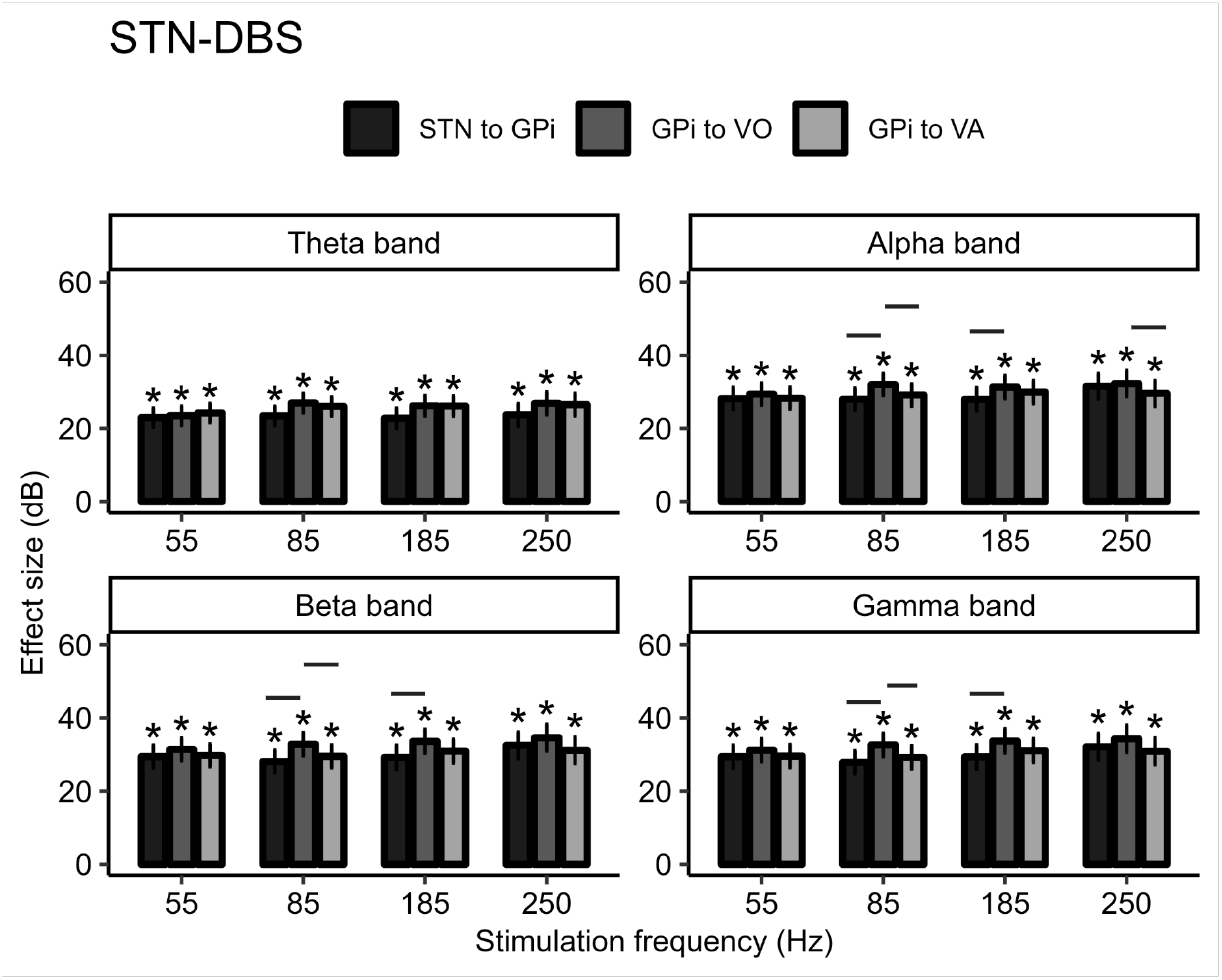
Statistical results: Effect of STN-DBS on the transfer function gains within the pathways.

#### Alpha band

The results of pairwise comparison with respect to baseline showed that during 85 Hz STN stimulation, GPi-VoaVop alpha band gain is significantly higher than that of STN-GPi (Estimate = 4.01: *p − value <* 0.01) and GPi-VA (Estimate = 3,36: *p − value <* 0.01). In addition, during 185 Hz STN stimulation, GPi-VoaVop alpha band gain is significantly higher than that of STN-GPi (Estimate = 3.37: *p − value <* 0.01) but not GPi-VA, which could mean that it does not interfere with GPi-VA transmission. STN-DBS at 250 Hz resulted in increased GPi-VoaVop versus GPi-VA transmission (Figure 7).

#### Beta band

The results of pairwise comparison with respect to baseline showed that, similar to alpha band, during 85 Hz STN stimulation, GPi-VoaVop alpha band gain is significantly higher than that of STN-GPi (Estimate = 3.75: *p−value <* 0.01) and GPi-VA (Estimate = 2.79: *p−value <* 0.01). In addition, during 185 Hz STN stimulation, GPi-VoaVop alpha band gain is significantly higher than that of STN-GPi (Estimate = 3.40: *p − value <* 0.01) but not GPi-VA. However no significant changes were observed within pathways due to 250 Hz STN-DBS (Figure 7).

#### Gamma band

The results of pairwise comparison with respect to baseline showed that, similar to alpha and beta band, during 85 Hz STN stimulation, GPi-VoaVop alpha band gain is significantly higher than that of STN-GPi (Estimate = 3.83: *p − value <* 0.01) and GPi-VA (Estimate = 2.85: *p − value <* 0.01). In addition, during 185 Hz STN stimulation, GPi-VoaVop alpha band gain is significantly higher than that of STN-GPi (Estimate = 3.41: *p − value <* 0.01) but not GPi-VA (Figure 7).

In summary, as shown in Figure 7, the GPi-VoaVop gains at the commonly used clinical stimulation frequencies for STN-DBS (85 and 185 Hz) are significantly larger than those of STN-GPi in alpha, beta, and gamma bands, but not the theta band. Moreover, the statistical analysis on the effect of STN-DBS on the transfer function gains at each frequency band is shown in Figure 8. These results illustrate that the STN-DBS has the lowest effect size (least increase) on the theta band transmission and the highest at beta and gamma band transmission. Meaning that the DBS facilitates transmission of beta and gamma more than the transmission of theta activity in pallido-thalamic network.

**Fig. 8.**
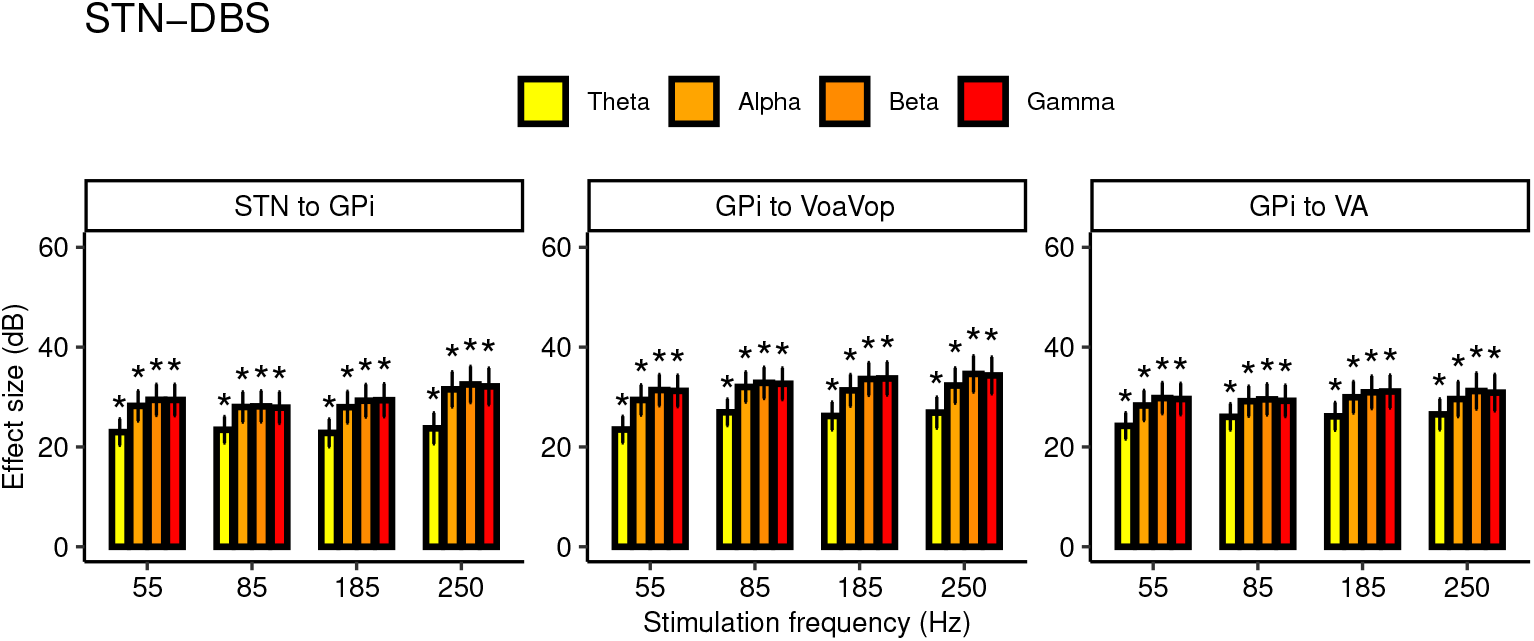
Statistical results: Effect of STN-DBS on the transfer function gains within the frequency bands for each pathway.

#### Clinically optimal versus non-optimal STN-DBS setting

Similarly, we marked the optimal stimulation setting for those patients that responded to STN-DBS, and this time, instead of using stimulation frequency as a predictor we tested whether the “optimal” and “non-optimal” stimulation settings are different from each other and baseline. We only had 3 patients who responded to STN-DBS. We used the same linear mixed effects model equation (Eq. 4) to fit the models. After the models were fit for each frequency band gain 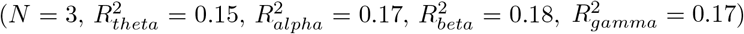, we performed a pairwise comparison to determine whether the “optimal” and “non-optimal” settings had different effects on the transmission of signals within deep brain regions. Figure 9 shows all the transfer function gains in four frequency bands (theta, alpha, beta, gamma), for all pathways. Each color represents one patient with their mean gain and standard deviation. No significant effect associated with the best clinical setting was observed due to STN-DBS; however, we cannot draw a definitive conclusion from these observations due to the small effect size and limited number of patients responsive to STN-DBS. For simplicity of the figure we are not showing the significance between the DBS-off versus optimal and non-optimal stimulation setting.

**Fig. 9.**
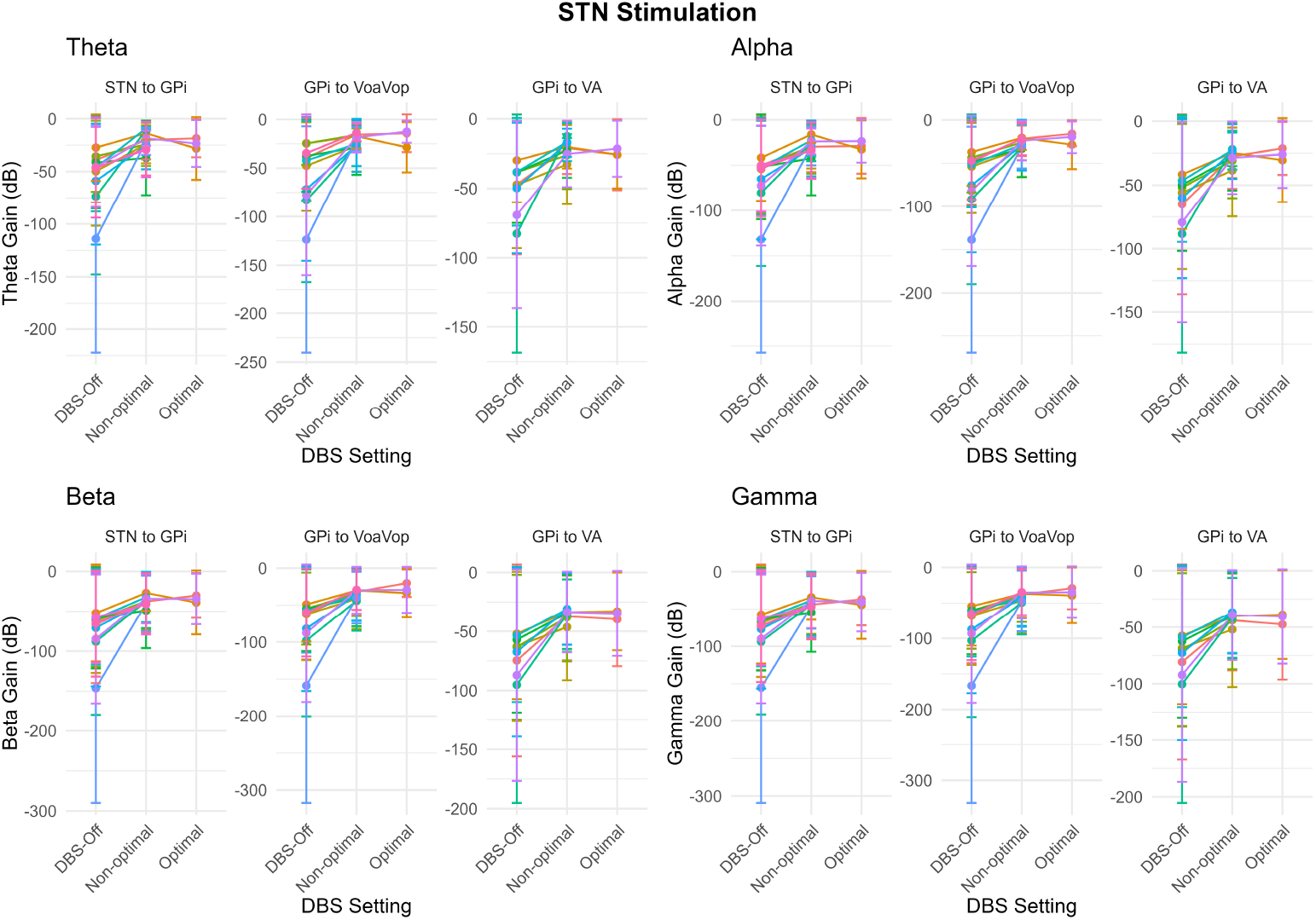
Effect of optimal versus non-optimal STN-DBS setting of transfer function gains, shown for each pathway and frequency band, for all patients.

## 4 Discussion

In this study, we demonstrate that DBS modulates physiological signal transmission within the basal ganglia-thalamic circuitry in children and young adults with dystonia. Using a novel transfer function analysis applied to intracranial recordings, we show that DBS—particularly GPi stimulation—alters the flow of frequency-specific neural activity across key motor pathways. The work done in this study suggests that GPi-DBS and STN-DBS affect the physiological connectivity and transmission patterns between deep brain areas in the motor circuitry, similar to the patterns of evoked responses across deep brain regions [47–49]. As such, one possible mechanism of DBS could be to restore balance between direct and indirect pathway gains by modulating activity propagating from the pallidum to the thalamus, either through suppression or facilitation of these pathways.

GPi-DBS affects neural transmission from STN to GPi, GPi to VoaVoa, and GPi to VA, such that the transfer function gains during all DBS settings presented in this work increase significantly from baseline. Our results show that GPi-DBS increases the GPi-VoaVop gain more than it affects the other two pathways (STN to GPi and GPi to VA) in beta and gamma bands, but not in theta and alpha bands. This observation supports the hypothesis that GPi-DBS may work by jamming the pallido-thalamic projection (GPi-VoaVop) by facilitating the transmission of higher frequency activity and “less-facillitating” the transmission of lower frequencies, as supported by Figure 5. However, no significant pattern of effects of the stimulation frequencies were observed on these gains during GPi-DBS.

To summarize, during GPi-DBS, given the excitatory output of STN and the inhibitory output of GPi, the observed reduction in STN-GPi transfer function gain relative to the GPi-VoaVop gain may imply: 1) GPi-DBS may reduce the overall excitability of GPi compared to other regions, resulting in disinhibition of thalamic subnuclei (VoaVop and VA). 2) GPi-DBS may disrupt the effective transmission of neural activity from GPi to thalamic targets by introducing noise and thereby reducing the signal-to-noise ratio, rather than selectively suppressing pathological signal projection associated with dystonia (e.g., theta band). 3) Reduced GPi activity or antidromically induced inhibition of STN could attenuate inhibitory drive to thalamic nuclei, thereby facilitating increased thalamic activity [50].

In addition, results from STN-DBS at 85 and 185 Hz, which are typically considered clinically effective stimulation frequenciesfor STN, showed a significant increase in GPi-VoaVop gains in higher frequency bands, reflecting increased amplification of alpha, beta, and gamma signals from GPi to VoaVop. This could indicate that STN-DBS has a more significant effect on downstream targets, potentially by decreasing GPi excitability and thereby increasing the VoaVop activity in those specific frequency bands. The group analysis and the analysis of the optimized clinical setting, including comparisons across clinical stimulation settings, did not reveal any significant effect of STN-DBS on theta band activity. In addition, the fact that we did not observe significant changes in LFO transmission within the pallido-thalamic network due to 55 and 250 Hz STN-DBS provides support for their therapeutic ineffectiveness in pediatric dystonia. Another important feature of the STN-DBS findings is facilitation of increased alpha, beta, and gamma transmission across all examined nuclei, relative to theta transmission. This may suggest that increased transmission of disorder-irrelevant activities (alpha, beta, and gamma) could interfere with the propagation of dystonia-related signals, particularly in the theta frequency range.

The limitations of this study include the heterogeneity of our patient cohort in terms of symptomatology, underlying disease etiology, and their treatment approaches (e.g. not all subjects had VA lead, and not all had VoaVop recordings), which contributed to variability in the effects of DBS across regions. Nevertheless, this heterogeneity also supports the broader applicability of the technique across diverse clinical populations. The use of data from micro-contacts is unique in this study, as most recorded information from DBS is either available from intraoperative microelectrode recordings, or postoperative macro-contact LFPs. Micro-contacts are likely to sample from smaller target regions than macro-contacts and thus may provide more specificity to the analysis of information transmission, though this comes with the trade-off of reduced spatial coverage. Comparison of this technique between micro-contacts and macro-contacts is an important topic for future work.

Moreover, it is important to consider that the methodology relies on the assumption of linearity in the system which may not hold true in all cases. The presence of transmitted information between two regions in the control system model does not necessarily imply that neural signals are transmitted via a direct anatomical connection. For example, the observed transmission, reflected by the transfer function gains, could arise from a common input projecting to both nuclei rather than true directed transmission. However, the regularization parameter applied during transfer function estimation accounts for additional inputs to the output nucleus and mitigates the influence of such indirect effects, thereby allowing us to extract meaningful insights from the computed transfer functions.

Another limitation lies in the presence of low-frequency noise, especially near the filter cutoff frequency (1 Hz), which may have compromised the reliability of results in the delta band compared to higher frequency bands. Furthermore, it is possible that important physiological information exists outside the analyzed frequency range (1–50 Hz), which was not captured in the current analysis.

In our method for calculation of the transfer functions we only calculate the signal amplification by computing the spectrum and magnitude of cross-spectrum of two signals; therefore, we do not take phase information into account. This approach is appropriate to our hypothesis which focuses on total information transmission and amplification between regions which is most affected by the transfer function gains. However, we acknowledge that the transfer function phase is important and contain very useful information about how the stimulation affects the phase and signal transmission delays; since it is highly possible that DBS introduces not only amplitude changes but also phase shifts that could affect the pattern of neural transmission. This will be another important topic for future analysis.

In conclusion, our findings provide evidence that DBS modulates the transmission of neural signals across basal ganglia-thalamic pathways, particularly enhancing low-frequency transmission from GPi to thalamic motor nuclei. This supports the notion that DBS may exert its therapeutic effects by disrupting abnormal inhibitory signaling patterns implicated in dystonia. These results not only offer mechanistic insight into DBS function but also underscore the potential utility of transfer function analysis in guiding future developments in closed-loop neuromodulation and adaptive neurotechnology.

## Data Availability

All data produced in the present study are available upon reasonable request to the authors. (Confidential data)

## Acknowledgements

We thank our volunteers and their parents for participating in this study. We also thank Jennifer MacLean and Teresa Serna for their assistance with neurological examinations and Alireza Seyyed Mousavi for helping in data collection. Moreover, we thank Dr. Joffre E. Olaya and Dr. Mark for performing the DBS surgeries. This study is funded by the Cerebral Palsy Alliance Research Foundation (PG02518).

## Authors Contributions

M.K., M.A., J.N. and T.D.S. conducted the experiments and collected the data. M.K. analyzed the data and performed statistical analysis. S.A. performed the image analysis and electrode localization. M.K. and T.D.S. interpreted the results of the study. J.O. and M.L. conducted the DBS surgeries. T.D.S. performed the neurological examination. M.K., M.A., J.N. and T.D.S. wrote the manuscript. All authors contributed to the final manuscript.

## 5 Supplementary Materials

### 5.1 Additional analysis: DBS modulates the oscillations associated with dystonic signals in deep brain regions

Previous studies of dystonic patients show abnormalities in low frequency activity in GPi and other motor sensory regions, such as STN, VoaVop and VA nuclei of thalamus. In this chapter we showed that DBS works in part by altering transmission of abnormal signals in low frequency bands between different brain regions. Here we want to examine and show that this effect can be both at the stimulation site (locally) and across deep brain regions (distant regions). In other words, we hypothesize that DBS modulates the abnormal projections onto thalamic motor subnuclei by changing the pattern of transmission in pallido-thalamic network, as a result of local and global changes in deep brain regions low frequency oscillations. To test this hypothesis, we performed a similar analysis on the power spectra of each region with and without the DBS. Furthermore, to evaluate the differences between the optimal versus non-optimal DBS settings for our patients, we analyzed the effect of clinically effective DBS on the PSDs and compared that with the effect of non-optimal DBS. The results from the PSD analysis confirms an increase and decrease in power that are both local and global in all regions. We confirmed the presented results (increased transmission from GPi to VoaVop) by showing that while stimulating in GPi, the GPi power effectively decreases and the VoaVop and VA powers increase significantly from baseline, resulting in increased transfer function gains from GPi to thalamic subnuclei. This also can be confirmed by the known fact that GPi projections onto thalamic nuclei is inhibitory, therefore decreased GPi activity essentially leads to increased VoaVop activity and increased transfer function gain. The group analysis on the clinically effective setting showed that the clinical DBS decreases the oscillations in all regions. The optimal GPi DBS showed significantly smaller VA, VoaVop, and STN activity in all bands versus non-optimal setting. However, GPi-DBS in optimal setting led to a significantly higher GPi power in theta and alpha bands compared to the non-optimal setting, which again confirms our results from the transfer function study. These results elicit a better understanding of the mechanism and effects of DBS.

### Footnotes

5 “Estimates” are the estimates of the effect sizes.

---

**Fig. 10.**
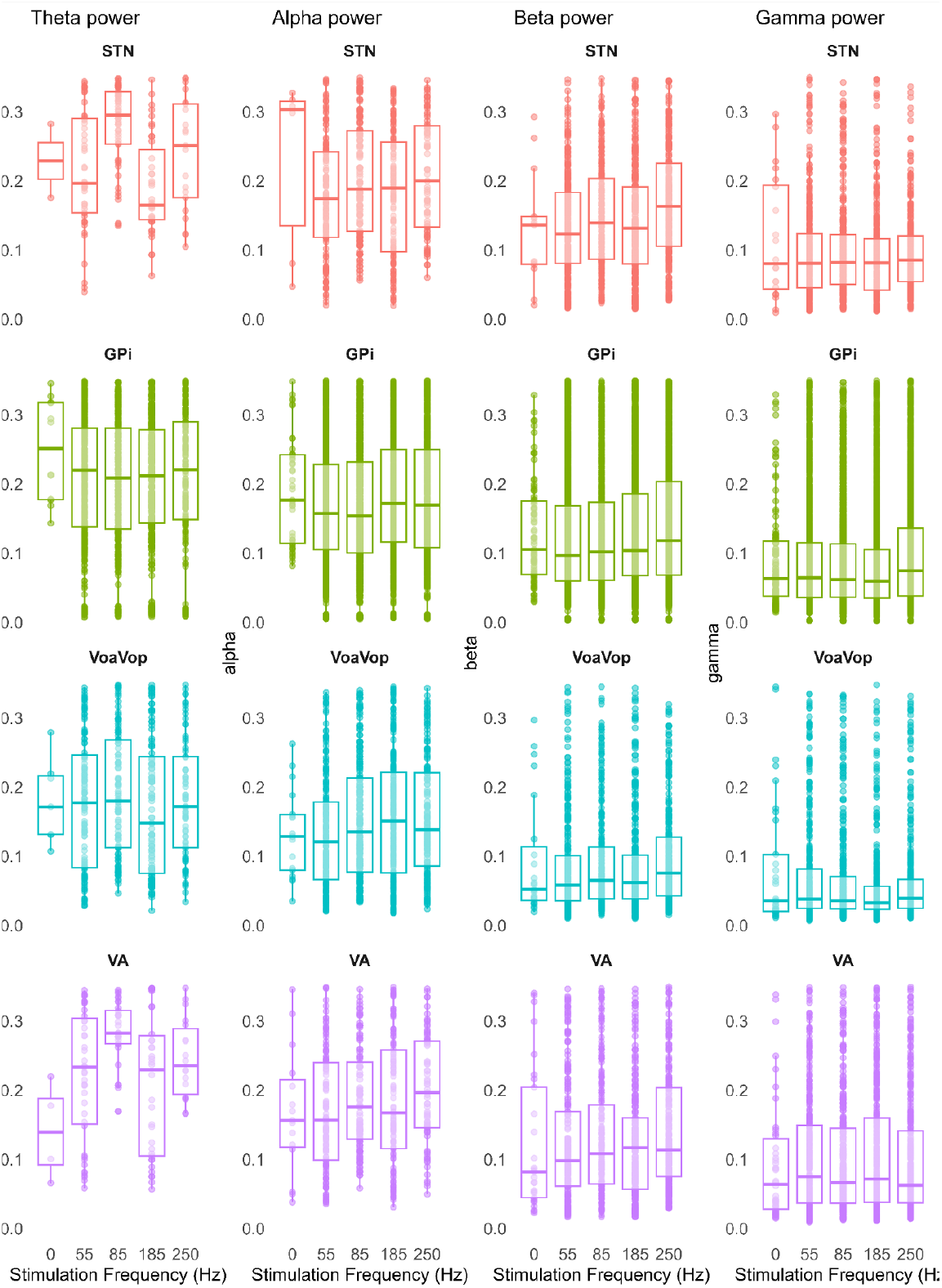
GPi stimulation: stimulation frequency effect on theta, alpha, beta, and gamma band powers in STN, GPi, VoaVop, and VA.

**Fig. 11.**
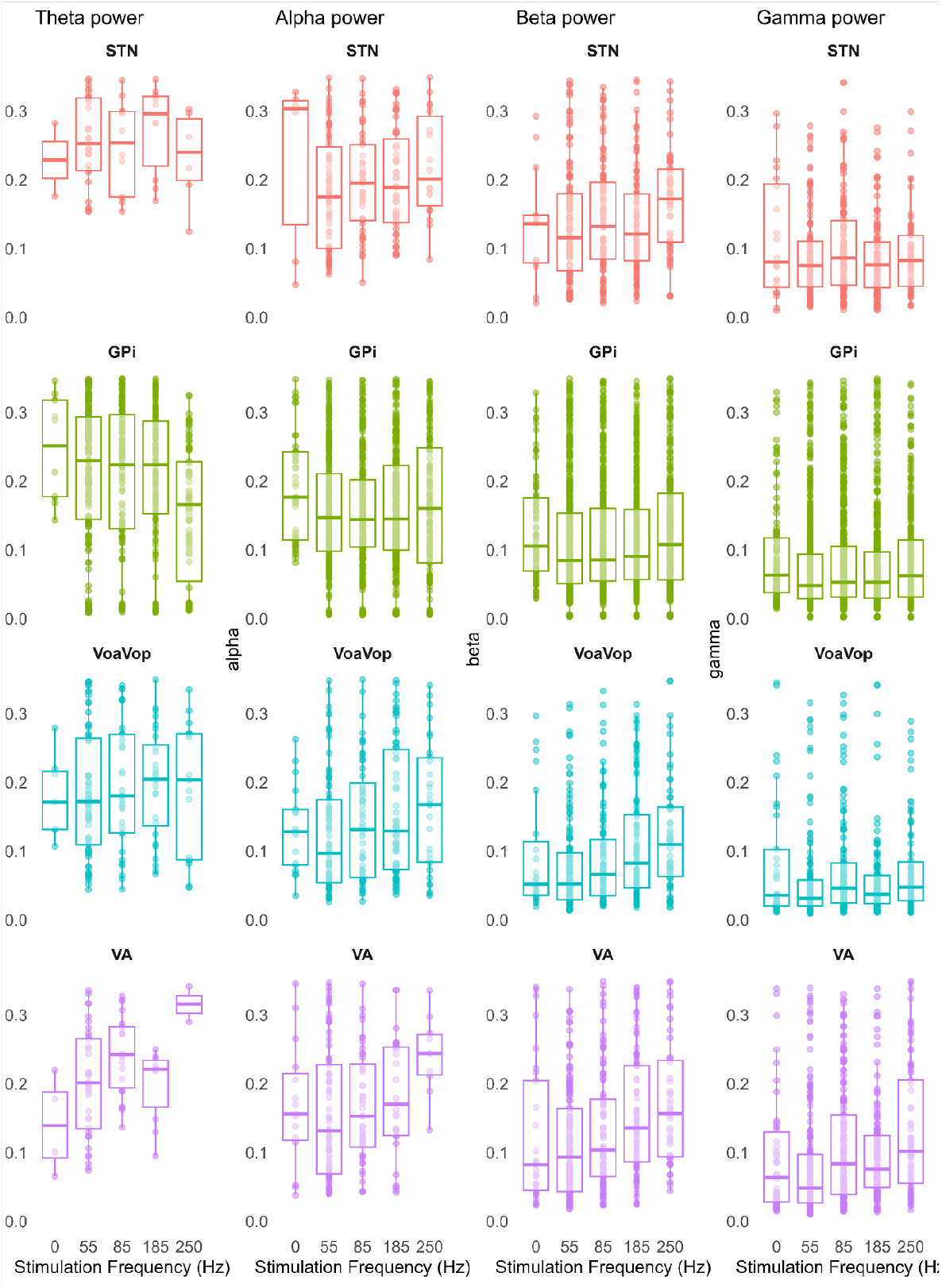
STN stimulation: stimulation frequency effect on theta, alpha, beta, and gamma band powers in STN, GPi, VoaVop, and VA.

**Fig. 12.**
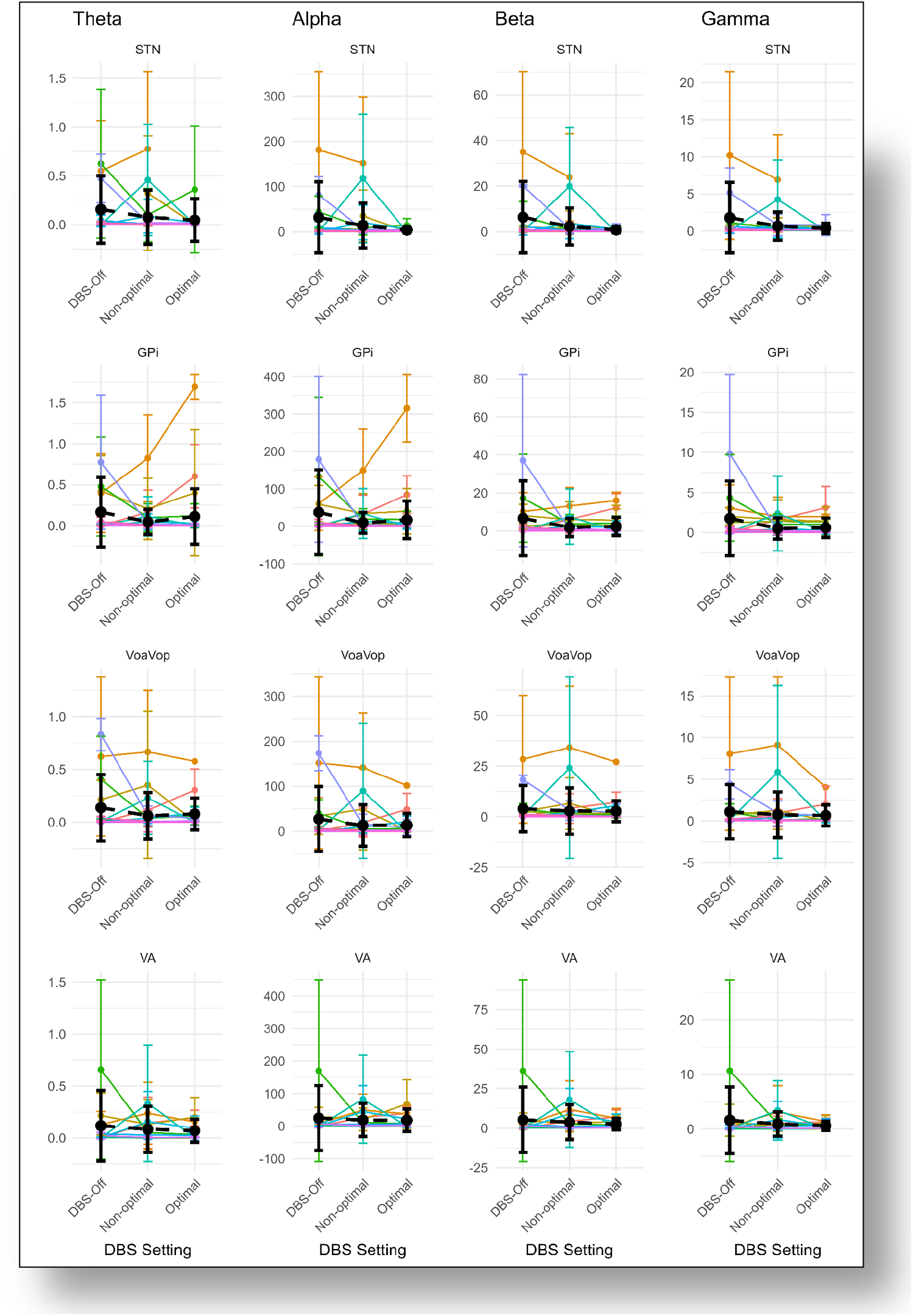
GPi stimulation: DBS-off, non-optimal, and optimal stimulation setting (stimulation frequency, stimulation lead contact and location): in theta, alpha, beta, and gamma bands (N=13)

**Fig. 13.**
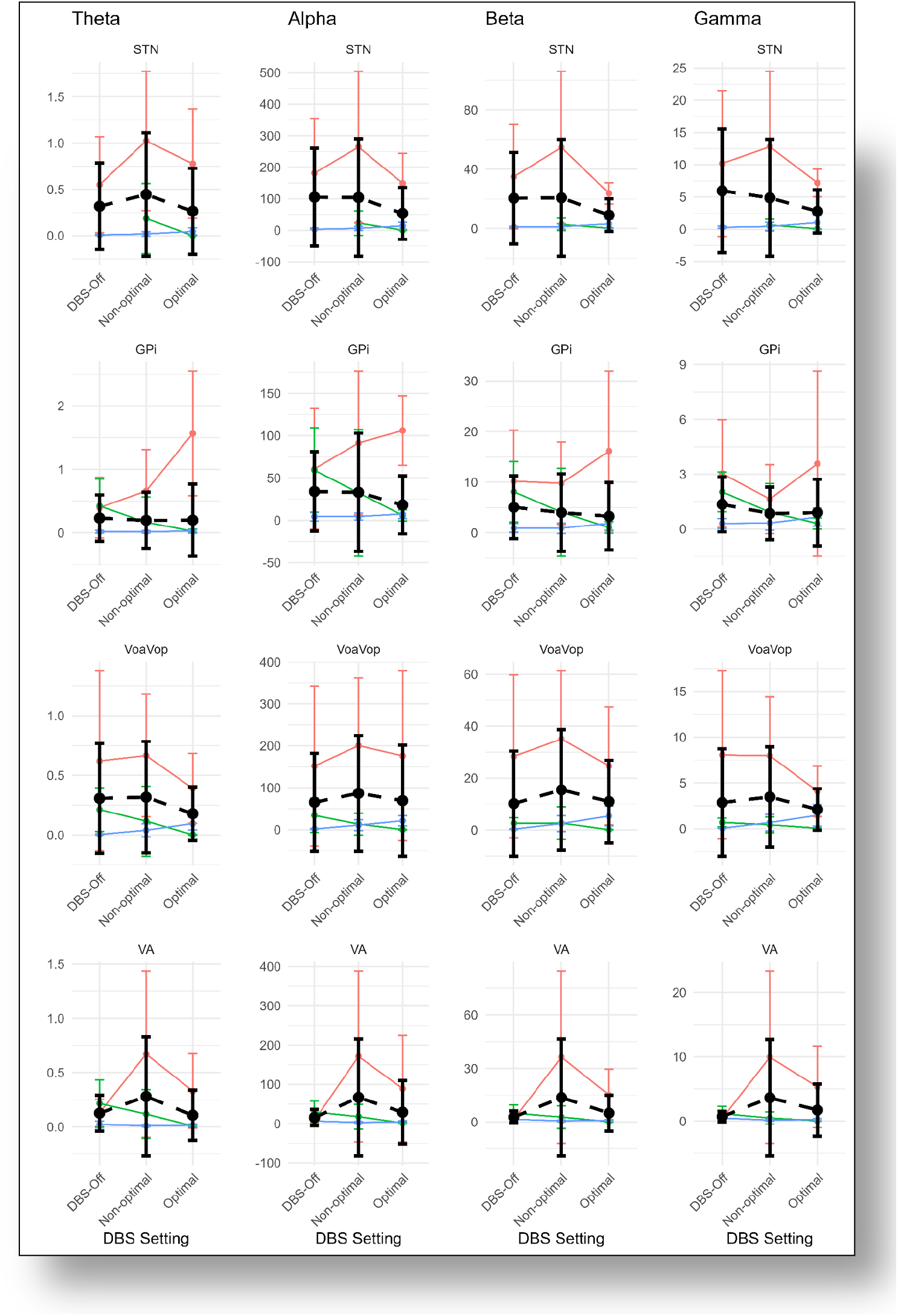
STN stimulation: DBS-off, non-optimal, and optimal stimulation setting (stimulation frequency, stimulation lead contact and location): in theta, alpha, beta, and gamma bands (N=3)

